# Defining the nuclear genetic architecture of a common maternally inherited mitochondrial disorder

**DOI:** 10.1101/2022.11.18.22282450

**Authors:** Róisín M. Boggan, Yi Shiau Ng, Imogen G. Franklin, Charlotte L. Alston, Emma L. Blakely, Boriana Büchner, Enrico Bugiardini, Kevin Colclough, Catherine Feeney, Michael G. Hanna, Andrew T. Hattersley, Thomas Klopstock, Cornelia Kornblum, Michelangelo Mancuso, Kashyap A. Patel, Robert D. S. Pitceathly, Chiara Pizzamiglio, Holger Prokisch, Jochen Schäfer, Andrew M. Schaefer, Maggie H. Shepherd, Annemarie Thaele, Rhys H Thomas, Doug M. Turnbull, Cathy E. Woodward, Gráinne S. Gorman, Robert McFarland, Robert W. Taylor, Heather J. Cordell, Sarah J. Pickett

## Abstract

Maternally inherited mitochondrial diseases are caused by pathogenic mitochondrial (mt)DNA variants. Affecting individuals at any age, they are often multi-systemic and manifest extreme clinical variability. We have limited understanding of the cause of this heterogeneity, which makes disease diagnosis and prognosis exceptionally challenging. This is clearly demonstrated by disease caused by m.3243A>G, the most common pathogenic mtDNA variant. m.3243A>G can cause a severe syndrome characterised by mitochondrial encephalomyopathy lactic acidosis and stroke-like episodes (MELAS), but individuals who carry m.3243A>G may be asymptomatic or manifest with any number of a range of phenotypes. There is strong evidence for the presence of nuclear factors that modify phenotype; we set out to characterise the nature of this nuclear involvement using genetic linkage analysis.

We assembled a multi-centre cohort of well-characterised patients and their maternal relatives, comprising 76 pedigrees, and characterised the nuclear genetic landscape of m.3243A>G-related disease phenotypes using non-parametric genetic linkage analysis. We considered eight of the most common m.3243A>G-related phenotypes, accounted for known risk factors using logistic regression, and determined empirical significance using simulation to identify regions of the nuclear genome most likely to contain disease modifying variants.

We identified significant genetic linkage to encephalopathy on chromosome 7q22, and suggestive regions for encephalopathy, stroke-like episodes and psychiatric involvement on chromosomes 1, 5, 6, 11 and 13. These findings suggest that these neurological features are likely to be influenced by a small number of nuclear factors with a relatively large effect size. In contrast, no linkage regions were identified for cerebellar ataxia, migraine, diabetes mellitus, hearing impairment or chronic progressive external ophthalmoplegia.

The genetic architecture of the nuclear factors influencing disease related to m.3243A>G differs between phenotypes. Severe and cardinal neurological features of MELAS are likely to be strongly influenced by a small number of nuclear genes, whereas the nuclear influence over other phenotypic presentations is more likely to be polygenic and complex in nature, composed of a larger number of factors that each exert a small effect. These results will inform strategies for future studies to identify the genes and pathways that influence clinical heterogeneity in m.3243A>G-related disease, with the ultimate aim of better understanding disease development and progression.

## Introduction

Maternally inherited mitochondrial DNA (mtDNA) disorders affect ∼1 in 5000 individuals, can arise at any age, are often multi-systemic and are extremely clinically variable^1,2^. The cause of this extreme heterogeneity, even within groups of individuals carrying the same pathogenic mtDNA variant, is poorly understood, making disease diagnosis and prognosis exceptionally challenging. This is exemplified by disease associated with the most common pathogenic mtDNA variant, m.3243A>G (RefSeq:NC_012920.1), within *MT-TL1*. Although initially identified as a cause of mitochondrial encephalomyopathy, lactic acidosis, and stroke-like episodes (MELAS)^3,4^, this syndrome only affects ∼15% of all m.3243A>G disease patients. Individuals can present with any number of a broad spectrum of clinical features including severe CNS phenotypes such as encephalopathy, stroke-like episodes, seizures, and cerebellar ataxia, as well as features outside of the CNS such as myopathy, cardiomyopathy, chronic progressive external ophthalmoplegia (CPEO), diabetes mellitus, renal impairment and sensorineural hearing loss. Estimates of the carrier frequency of m.3243A>G are 40-70 times higher than disease prevalence, suggesting that many individuals who carry m.3243A>G are clinically asymptomatic or have a phenotype not recognised as mitochondrial^1,5^.

A contributing factor to mtDNA disease complexity is the co-existence of both wild-type and mutant mtDNA molecules within the same cell (known as heteroplasmy); m.3243A>G levels can vary between individuals, tissues and cells^6–9^. Variation in tissue m.3243A>G heteroplasmy within commonly tested tissues and age both contribute to the heterogeneity of phenotypic presentation, disease severity and progression but the vast majority (∼70-80%) of variability remains unexplained^8–12^. Moreover, patients with similar m.3243A>G levels can present with very different phenotypes and disease severity, highlighting how difficult it is to provide patients with satisfactory genetic counselling.

The mitochondrial genome encodes 13 protein-coding genes, 2 rRNAs, and 22 tRNAs: all essential to mitochondrial function^13^. In contrast, the nuclear genome encodes ∼1,200 proteins necessary for mitochondrial function^14^; pathogenic variants in either genome can result in mitochondrial disease^2^. We have previously demonstrated that nuclear genetic variation contributes to the phenotypic diversity of m.3243A>G-related disease; moderate to high heritability estimates indicate that additive genetic factors play a much larger role in determining disease phenotype than m.3243A>G proportion, age, and sex combined^12^. However, we do not know whether these factors are monogenic or polygenic and the specific factors have yet to be identified.

With the aim of understanding the nuclear genetics contributing to this heritability, we assembled a unique multi-centre cohort of well-characterised patients and their maternal relatives, comprising 76 pedigrees. This enabled us to characterise the nuclear genetic landscape of m.3243A>G-related disease phenotypes using non-parametric genetic linkage analysis. This technique has been highly successful in identifying regions of the nuclear genome that co-segregate with numerous disease phenotypes (e.g. type 1 diabetes^15^, multiple sclerosis^16^, Crohn’s disease^17^) and has the advantage of being able to detect effects due to rare and common variants, along with family-specific effects^18,19^. We considered eight of the most common m.3243A>G disease-related phenotypes and accounted for known risk factors by using the residuals from logistic regression, identifying regions of the nuclear genome most likely to contain disease modifying variants.

## Methods

### Study cohort

Our combined cohort consists of 488 individuals from five clinical centres (**Table 1**; 192 male; median age=41.7, IQR=22.9), all genetically confirmed to carry the m.3243A>G variant. Written informed consent from patients was obtained prior to study inclusion and all clinical investigations were evaluated according to the Declaration of Helsinki.

**Table 1.** Demographics of cohorts that were combined to create a cohort of 488 m.3243A>G variant carriers. *Individuals from Exeter originate from a specialist diabetes clinic, 110 individuals have phenotypic information for diabetes, the number of individuals with documented assessment for other phenotypes range from 5-18.

| **Cohort** | **Number of individuals with phenotype assessment (M, F)** | **Median age at last assessment (IQR, range)** | **Median m.3243A>G level (IQR, range)** |
| --- | --- | --- | --- |
| **Newcastle, UK** | 251<br>(103, 148) | 45.8<br>(24.1, 18.1-79.9) | 0.648<br>(0.556, 0.002-1.000) |
| **Germany** | 56<br>(25, 31) | 45.0<br>(20.5, 22.0-80.0) | 0.637<br>(0.465, 0.058-1.000) |
| **Italy** | 10<br>(6, 4) | 45.0<br>(9.75, 33.0-58.0) | 0.696<br>(0.169, 0.490-1.000) |
| **University College London (UCL), UK** | 56<br>(22, 34) | 44.5<br>(25.8, 19.0-79.0) | 0.938<br>(0.394, 0.074-1.000) |
| **Exeter, UK** | 115*<br>(36, 79) | 34.0<br>(17.0, 5.0-69.2) | 0.816<br>(0.324, 0.002-1.000) |
| **Combined** | 488<br>(192, 296) | 41.7<br>(22.9, 5.0-80.0) | 0.743<br>(0.053, 0.002-1.000) |

There were 251 individuals included from the Newcastle component of the Mitochondrial Disease Patient Cohort, UK, all of whom were reviewed and investigated by the NHS Highly Specialised Service for Rare Mitochondrial Disorders of Adults and Children in Newcastle upon Tyne. Ethical approval was granted by the Newcastle and North Tyneside Research Ethics Committee (13/NE/0326). Mitochondrial disease patient tissue was provided by the Newcastle Mitochondrial Research Biobank (16/NE/0267).

Fifty-four individuals were included from the University College London (UCL) arm of the Mitochondrial Disease Patient Cohort, UK (13/NE/0326); ethical approval was provided by the Queen Square Research Ethics Committee, London, UK (09/H0716/76).

Data and 56 samples were received from the German network for mitochondrial disorders “mitoNET”. The clinical Registry (mitoREGISTRY) was approved by the Ethics Committee of the LMU Munich (182-09) and local Ethic Committees of all contributing medical institutions. The Biobank (mitoSAMPLE) was approved by the Ethics committee of the Technical University Munich (200/15 S-SR) and local Ethic Committees of all contributing medical institutions.

We also included 115 individuals who were referred from routine clinical practice to the Exeter Genomics Laboratory at the Royal Devon and Exeter Hospital for diagnostic genetic screening related to diabetes mellitus and were shown to have m.3243A>G-related disease. Informed consent was obtained from all participants, and the study was approved by the North Wales ethics committee (17/WA/0327). Genetic testing revealed that six of these individuals were also in the Newcastle component of the Mitochondrial Disease Patient Cohort, UK. In these cases, phenotype data from both cohorts were merged into one record.

Ten individuals were included from the University of Pisa, enrolled in the “Nationwide Italian Collaborative Network of Mitochondrial Diseases”. The database establishment (and its use for scientific purposes) was approved by the local Ethical Committees of the single centres, which obtained written informed consent from all patients or legal representatives.

### mtDNA molecular analyses

Where available, blood m.3243A>G levels were obtained from the recruiting clinical centres. Where these data were not available, m.3243A>G levels were quantified within DNA samples that had been extracted from whole blood by pyrosequencing on the Pyromark Q24 platform, as previously described and validated (test sensitivity >3% mutant mtDNA)^20^.

Blood levels of m.3243A>G decline with age and so were corrected using the previously published formula^9^:

Age-adjusted blood level = Blood level/0.977^(age+12)^

Where multiple blood m.3243A>G levels were available, the mean age-adjusted blood level was used. Where blood levels of m.3243A>G level were not available, urine m.3243A>G levels were used (n=18), correcting for sex using the previously published formulas^9^:

male adjusted urine level=logit^-1^((logit(urine level)/0.791)-0.625)

female adjusted urine level=logit^-1^((logit(urine level)/0.791)+0.608).

Where neither blood nor urine measures were available, we used assessments from skeletal muscle tissue (n=5); no correction was performed on measures from muscle tissue, as levels remain stable over time.

### Phenotype assessments

Assessments of disease phenotypes in the form of Newcastle Mitochondrial Disease Adult Scale (NMDAS) scores were available within the cohorts from Newcastle, Germany, and Italy. The NMDAS is a clinically validated scale used to assess the severity of mitochondrial disease in adult patients, consisting of 29 questions rated on an ordinal scale of 0-5 (0=unaffected, 5=severely affected)^21^. We considered 14 clinician-rated questions from sections II and III of the NMDAS covering both neurological and other systemic involvements associated with m.3243A>G-related disease. We also considered patient-assessed hearing from section I, as hearing loss is common in 3243A>G-related disease and this is the only NMDAS question that assesses this common 3243A>G-related phenotype. We excluded five phenotypes from sections II and III; pyramidal, extrapyramidal and visual acuity (due to low phenotype frequencies within the cohort); ptosis (as ptosis is a component of CPEO and evaluation of longitudinal ptosis data showed assessments to be subject to considerable inter-assessor variation); and cognition (as data were not available across the whole cohort, due to language-related constraints in administering this test). In-line with previous studies^12^, where multiple clinical assessments were available for an individual, we used the maximum NMDAS score and the youngest age at which that score was attained (or the age at most recent assessment if NMDAS=0).

We converted these semi-quantitative scores into binary assessments of phenotype affection status (0=unaffected/asymptomatic, 1=affected) using previously published thresholds, developed with clinical guidance (**Table 2**)^12^. For the cohorts from UCL and Exeter, full NMDAS assessments were unavailable, and therefore a clinician used these descriptors to retrospectively assign a binary score for each phenotype, using information available within medical notes. Cohort demographics are summarised in **Table 2**.

**Table 2.** Phenotypes considered for phenotype and genetic linkage analysis. Phenotyped cohort: cohort used for logistic regression analyses to account for the effect of established risk factors. Linkage cohort: phenotyped individuals within pedigrees with SNP genotype information available. The linkage cohort is smaller than the phenotyped cohort as not all individuals were members of pedigrees. For all phenotypes, the median number of SNP genotyped individuals per pedigree was 3 (IQR=ranges from 3-4 dependent on phenotype, range=3-9), except for diabetes (median =2, IQR=3, range=3-9). CPEO: chronic progressive external ophthalmoplegia.

| Phenotype | Definition | NMDAS threshold score | Phenotyped cohort | Linkage cohort |  |
| --- | --- | --- | --- | --- | --- |
|  |  |  | Total (Affected) | Total (Affected) | Number of informative pedigrees |
| **Psychiatric involvement** | Moderate to severe, warranting specialist treatment | 3 | 378 (76) | 139 (22) | 54 |
| **Cerebellar ataxia** | At least unable to maintain heel-toe walking or displaying unilateral dysmetria | 2 | 375 (145) | 155 (50) | 60 |
| **Migraine** | Four days per month or more | 5 | 377 (89) | 156 (37) | 61 |
| **Neuropathy** | At least sensory impairment (e.g., glove and stocking sensory loss) | 2 | 375 (44) | 155 (14) | 60 |
| **Dysphonia-dysarthria** | Showing at least mild impairment (clear impairment but easily understood) | 2 | 376 (62) | 154 (19) | 60 |
| **Seizures** | At least past history of epilepsy | 1 | 380 (104) | 141 (29) | 55 |
| **Encephalopathy** | At least past history of encephalopathy | 1 | 378 (80) | 156 (27) | 61 |
| **Stroke-like episodes** | At least transient focal sensory symptoms | 1 | 378 (82) | 156 (24) | 61 |
| **CPEO** | Abduction of at least one eye incomplete | 2 | 374 (62) | 154 (17) | 60 |
| **Hearing impairment** | At least moderate deafness (e.g. regularly requiring repetition), not fully corrected with hearing aid) | 3 | 392 (218) | 162 (79) | 62 |
| **Gastro-intestinal disturbance** | Dysmotility requiring admission or persistent and/or recurrent anorexia/vomiting/weight loss | 4 | 377 (43) | 154 (17) | 60 |
| **Myopathy** | Mild but clear proximal weakness in hip flexion and shoulder | 2 | 375 (139) | 154 (48) | 60 |
| **Diabetes mellitus** | At least impaired glucose tolerance (in absence of intercurrent illness) | 2 | 487 (274) | 198 (93) | 76 |
| **Cardiovascular involvement** | At least asymptomatic left ventricular hypertrophy on echo or nonsustained brady/tachyarrhythmia on ECG | 2 | 356 (126) | 144 (43) | 56 |

### Accounting for established influences of m.3243A>G-related disease

To determine the effects of age, m.3243A>G variant proportion and biological sex on the development of each phenotype, we performed a logistic regression in R (version 3.6.0) using the ‘glm()’ function with the binomial option in the ‘stats’ package^22^. Odds ratios were calculated from the exponentials of the coefficients estimated by a logistic regression model; they represent the odds that an individual will move from unaffected to affected given an increase in age (decade) or 10% m.3243A>G variant level. McFadden’s pseudo-R^2^ values^23^ were calculated using the following formula: 1-log(L_c_)/log(L_null_) were L_c_ is the maximum likelihood of the fitted model, and L_null_ is the maximum likelihood of the null. To account for the effects of these known risk factors in downstream linkage analysis, model residuals were extracted and used as a proxy phenotype. Pearson correlation coefficients were used to estimate correlations between phenotypes and p values Bonferroni corrected for 171 tests.

### SNP genotyping and quality control

417 DNA samples were available for SNP genotyping, which was performed using the UKB_WCSG array, known as the UK Biobank Axiom® Array, designed by the UK Biobank Array Design Group. Initial data quality control was performed in Axiom® Analysis Suite (AxAS: version 4.0.3.3) using internal Axiom® quality metrics (Fisher’s Linear Discriminant, FLD ≥4; Call rate ≥ 97; Heterozygous ratio offset ≥ 0; Homozygous ratio offset ≥ 0). Data for 645,115 SNPs were subsequently exported in PLINK format.

A second round of quality control (with SNP exclusion thresholds chosen to be specific for linkage analysis) was performed in PLINK (version 1.90b6.6)^24^ in accordance with recommendations^25^ and visualisation was performed in R. Nuclear SNPs were pruned by minor allele frequency (>0.35) and linkage disequilibrium (using the PLINK command ‘--indep 50 5 2’). SNPs were subsequently thinned, by selecting the two SNPs with the highest heterozygosity in each 2.4cM window using the program MapThin^26^, resulting in a final set of 8,214 autosomal SNPs for linkage analysis. 408 genotyped samples remained after the completion of quality control procedures.

### Linkage analysis

Logistic regression allowed us to account for known risk factors by using the residuals from these models as proxy-phenotypes in downstream linkage analysis. A variant of Haseman- Elston regression-based linkage analysis, which regresses estimated identity-by-descent (IBD) between relative pairs on the squared sums and squared differences of trait values of the pairs, was used. This approach should be relatively robust to non-normality of the trait of interest. Population parameters (mean, variance, heritability) were derived from the full population sample. Multipoint logarithm of the odds (LOD) scores for the presence of a quantitative trait locus (QTL) were estimated by Merlin-REGRESS^27^, as implemented in Merlin (v1.1.2)^28^.

### Simulation studies

To determine the ability of the data to detect significant regions of genetic linkage, we performed 1,000 gene-dropping simulations in Merlin-REGRESS using the ‘--simulate’ option, retaining the allele frequencies, marker positions, and missing genotype patterns of the data. Simulated data was analysed in an identical way to the actual data. Regions of suggestive and significant linkage were initially identified using the widely accepted LOD score thresholds of LOD ≥ 1.86 and LOD ≥ 3.3 respectively^29^. Linkage peaks were defined using a base pair (bp) window surrounding each maximum LOD score. The performance of several window sizes was tested against manual identification of linkage peaks (by visual inspection) within the results data; ±15Mb resulted in the best concordance with manual identification of linkage regions.

The distribution of empirical LOD scores under the null model differed considerably between phenotypes, therefore, applying the standard significance threshold of LOD ≥ 3.3 is not appropriate (**Figure 1**)^30^. For some phenotypes (e.g. stroke-like episodes), the distribution of simulated LOD scores was truncated at the upper end indicating lack of informativeness; phenotypes with a maximum simulated LOD score below our suggestive linkage threshold (LOD < 1.86; neuropathy, seizures, gastro-intestinal disturbance, myopathy and cardiovascular involvement) were removed from further analysis. This process left eight remaining phenotypes for inclusion in linkage analysis: psychiatric involvement, cerebellar ataxia, migraine, encephalopathy, stroke-like episodes, CPEO, hearing impairment, and diabetes mellitus.

**Figure 1.**
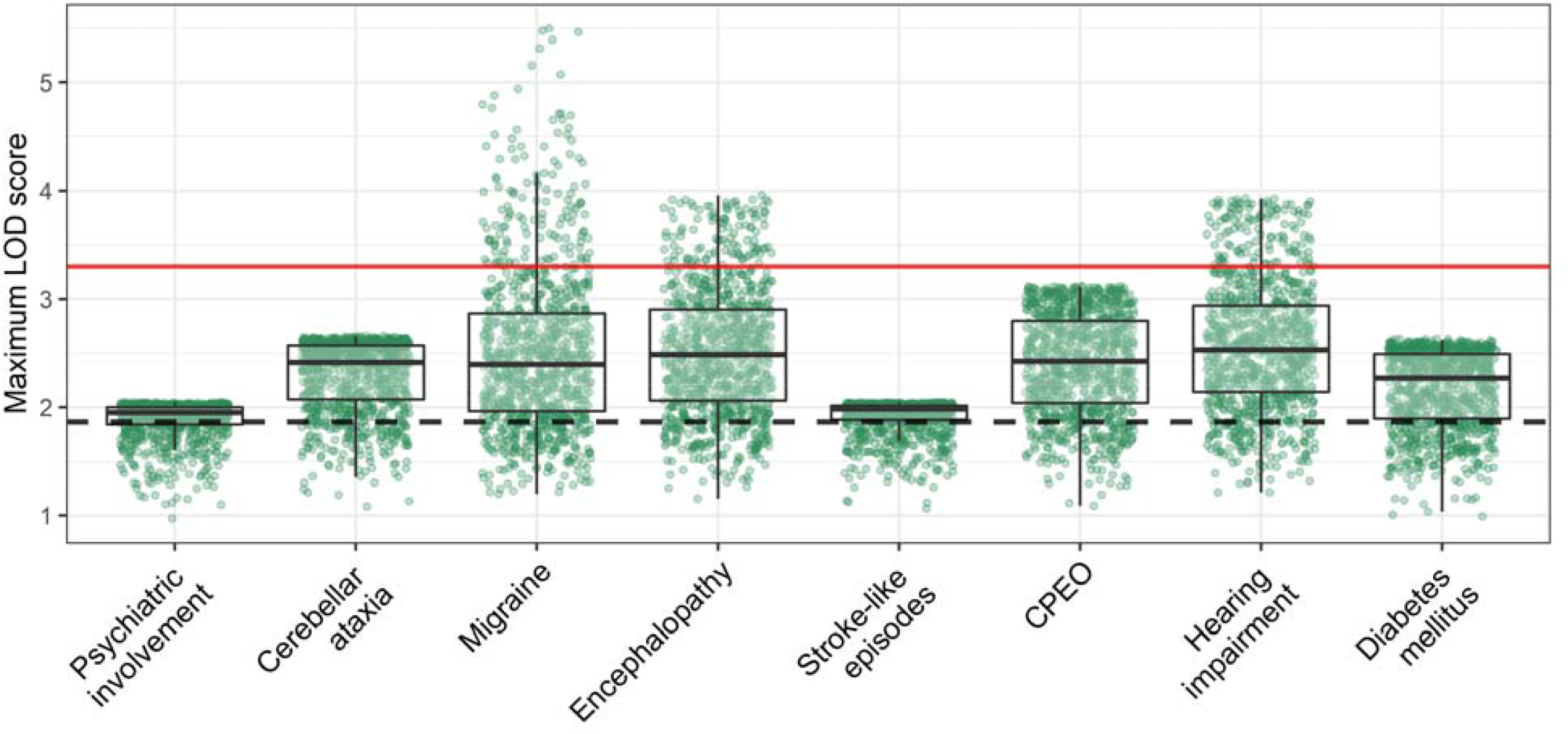
Maximum LOD scores achieved through simulation of genetic data. Each point represents the maximum LOD score per simulated genome scan, per phenotype. Nominal thresholds for significant (LOD ≥ 3.3; horizontal red line) and suggestive (LOD ≥ 1.86; horizontal dashed black line) linkage are shown for reference^29^. Phenotypes which demonstrated ‘topping out’ below LOD = 1.86 were removed from the analysis, as this indicated insufficient information was available for analysis. Box-plots depict the median, 25th and 75th quantiles and whiskers extend from the hinge to the largest/smallest values no further than 1.5 * IQR from the hinge.

Autosomal genome-wide empirical suggestive and significant LOD score thresholds for each phenotype were derived from simulated data (supplemental table 1). ‘Suggestive of genetic linkage’ was defined as ≥ the LOD score which appeared once per genome scan, and ‘significant genetic linkage’ as ≥ LOD score which appeared 0.05 times per genome scan (i.e once in 20 genome scans).

## Results

### Impact of age, m.3243A>G variant level, and biological sex on the development of m.3243A>G-related disease phenotypes

Previously, we established that m.3243A>G level (within commonly tested tissues) and age account for less than 20% of the variation in severity of m.3243A>G-related phenotypes. We replicated this finding in our expanded cohort using binary phenotype data (**Figure 2**), confirming that the proportion of variance explained is <20% for all phenotypes except psychiatric involvement and seizures (pseudo-R^2^=0.208 and 0.259, respectively). We also observed a sex effect in the analysis of encephalopathy (OR_male_=1.70, CI=1.01-2.85, *p=*0.004), CPEO (OR_male_=2.47, CI=1.15-3.40, *p*=0.019), and myopathy (OR_male_=0.59, CI=0.37-0.94, *p*=0.026). With the addition of biological sex as an explanatory variable, the proportion of observed phenotypic variation that can be explained for these select phenotypes is still <10%.

**Figure 2.**
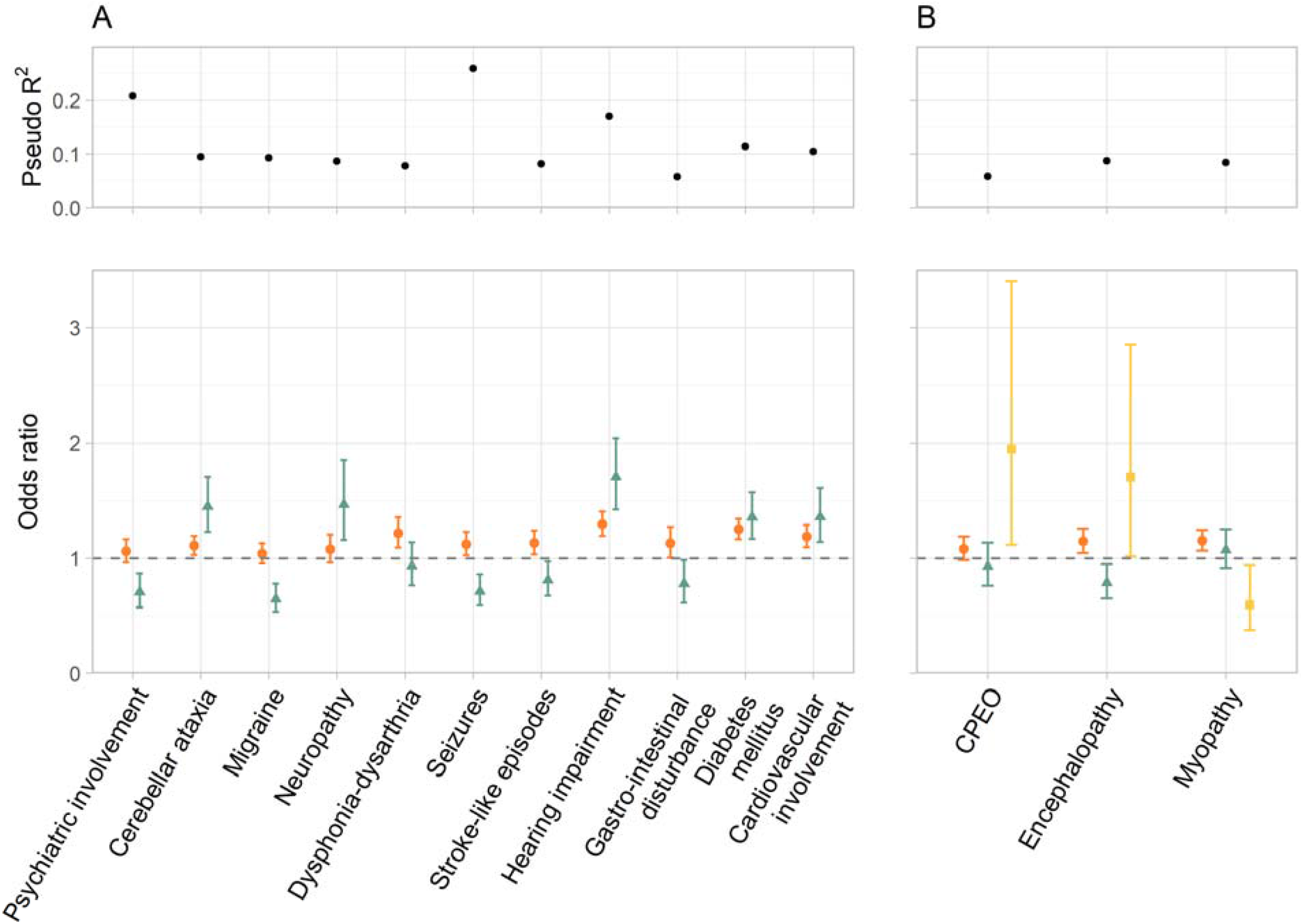
The proportion of phenotypic variation attributable to established factors. (A) Phenotypes in which the effect of sex was not significant; (B) phenotypes in which the effect of sex was significant. Reported are McFadden’s pseudo-R^2^ values and odds ratios (with 95% confidence intervals) for the effect of age in decades (green triangles), age-corrected m.3243A>G variant level in 10% increments (orange circles), and sex_male_ (yellow squares) on the development of m.3243A>G-related phenotypes (n=465 for diabetes, n=356-367 for all other phenotypes).

### Genetic linkage identified in neurological phenotypes of m.3243A>G-related disease

As these factors do not account for the majority of the phenotypic variation and nuclear modifiers are likely to contribute to this heterogeneity, we performed linkage analysis using the residuals from logistic regression as a quantitative phenotype and determined significance using phenotype-specific empirical suggestive and significant linkage LOD score boundaries (details in Materials and Methods).

Two of the most severe neurological phenotypes associated with m.3243A>G are stroke-like episodes, and encephalopathy. For encephalopathy, we identified regions suggestive of genetic linkage on chromosomes 1 (LOD_max_=2.24), 5 (LOD_max_=2.94), 11 (LOD_max_=3.48), and 13 (LOD_max_=2.61; **Figure 3A**), and a region of significant genetic linkage on chr 7 (LOD_max_=3.72; 7q22.3; **Figure 3B**). There are >150 genes within this region, 15 of which have a likely mitochondrial function as identified by MitoCarta^14^.

**Figure 3.**
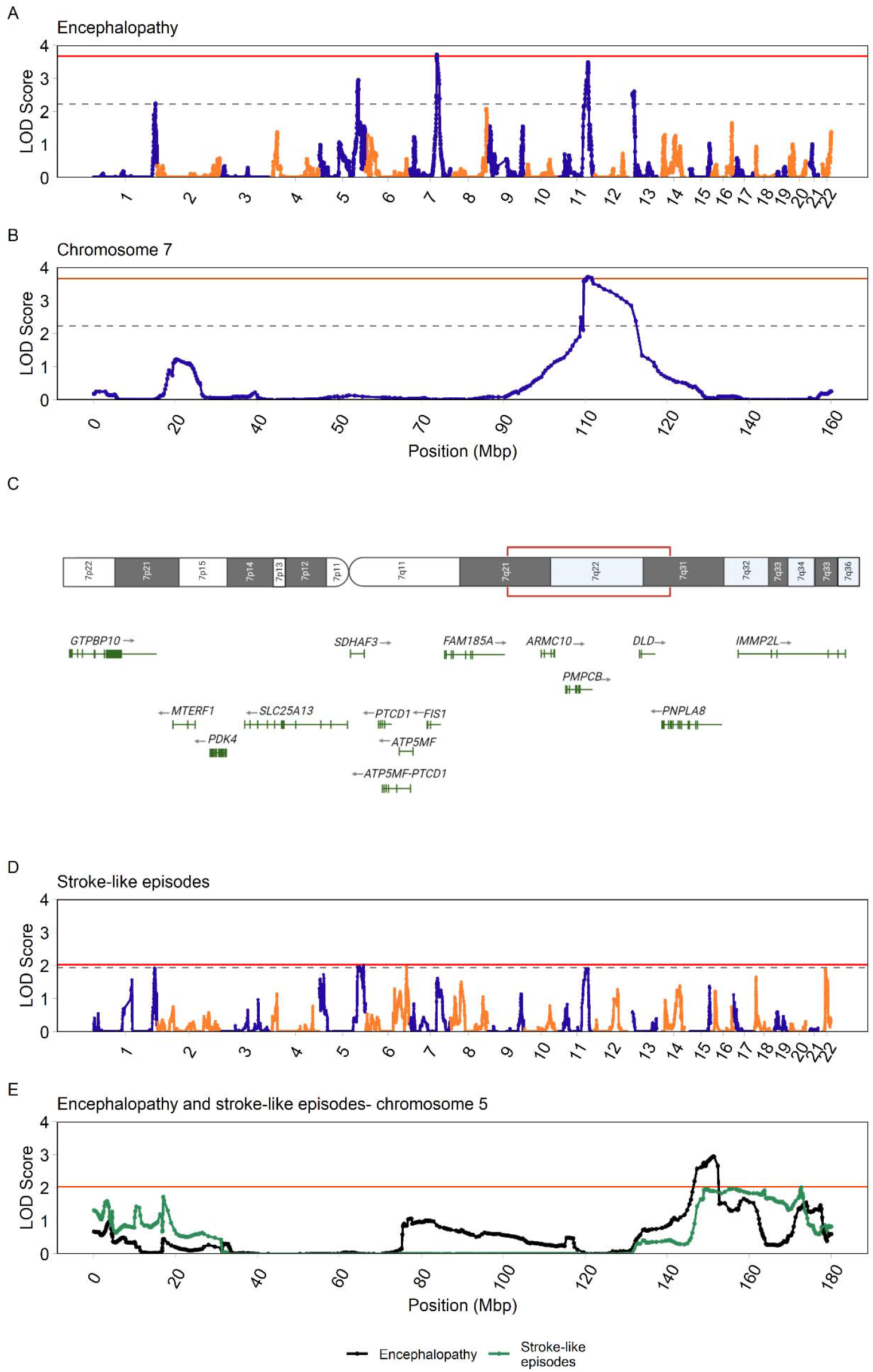
Linkage results for encephalopathy and comparison with results from stroke-like episodes. (A) Genome-wide linkage analysis of encephalopathy, empirical significance threshold LOD ≥ 3.67 (horizontal red line); empirical suggestive threshold LOD ≥ 2.23 (horizontal grey dashed line). A region on chromosome 7 (7q21.13-31.32) was identified as significant (LOD_max_=3.716) and a further four linkage regions were identified as suggestive of genetic linkage. (B) Encephalopathy chromosome 7 linkage. (C) Identified genes in the highlighted region on chromosome 7 have a likely mitochondrial function/localisation according to MitoCarta3.0^14^, positions and lengths of genes are relative, not to scale. (D) Stroke-like episodes, as recorded through the NMDAS assessment. Empirical suggestive threshold LOD ≥ 1.94, empirical significance threshold LOD ≥ 2.03. Two regions suggestive of genetic linkage were identified on chromosomes 5 (LOD_max_=2.01) and 6 (LOD_max_=1.96). (E) Comparison of chromosome 5 suggestive linkage regions for encephalopathy (LOD_max_=2.94), and stroke-like episodes (LOD_max_=2.01), stroke-like episodes LOD significance threshold (LOD ≥ 2.03) shown by red horizontal line.

For the stroke-like episodes phenotype, we identified two regions suggestive of genetic linkage on chromosome 5 (LOD_max_=2.01) and chromosome 6 (LOD_max_=1.96; **Figure 3C**); no regions reached our significance threshold of 2.03, although the pattern of LOD scores under the null suggests that our cohort has limited power to detect large LOD scores. As expected, there is some correlation between encephalopathy and stroke-like episodes, both cardinal features of MELAS, (r∼0.54, p<0.0001) and the region identified for stroke-like-episodes on chromosome 5 (apex at 5q33.2) overlaps with a region showing suggestive evidence of linkage to encephalopathy (apex at 5q33.1; **Figure 3D**); this may indicate a shared nuclear factor that influences both phenotypes.

Psychiatric involvement is relatively common in our cohort at ∼20%, particularly in combination with encephalopathy and stroke-like episodes (r∼0.12-0.27, p=0.0001-0.0193) and has been previously shown to be heritable^12^. The NMDAS assessment of psychiatric involvement covers a broad range of psychological presentations; to be classed as affected by a psychiatric disorder in this study, individuals must have an NMDAS score >=3, indicating that their level of psychiatric involvement is moderate to severe, requiring specialist treatment. A region of suggestive genetic linkage (LOD ≥ 1.89) was identified on chromosome 1 (LOD_max_=1.97; 1p36.22; **Figure 4A)**. Null-simulated maximum LOD scores demonstrate an upper threshold (**Figure 1**), which suggests that the information contained within the pedigrees used in the analysis is limited, and that an increased sample size is necessary to make informed conclusions from these results.

**Figure 4.**
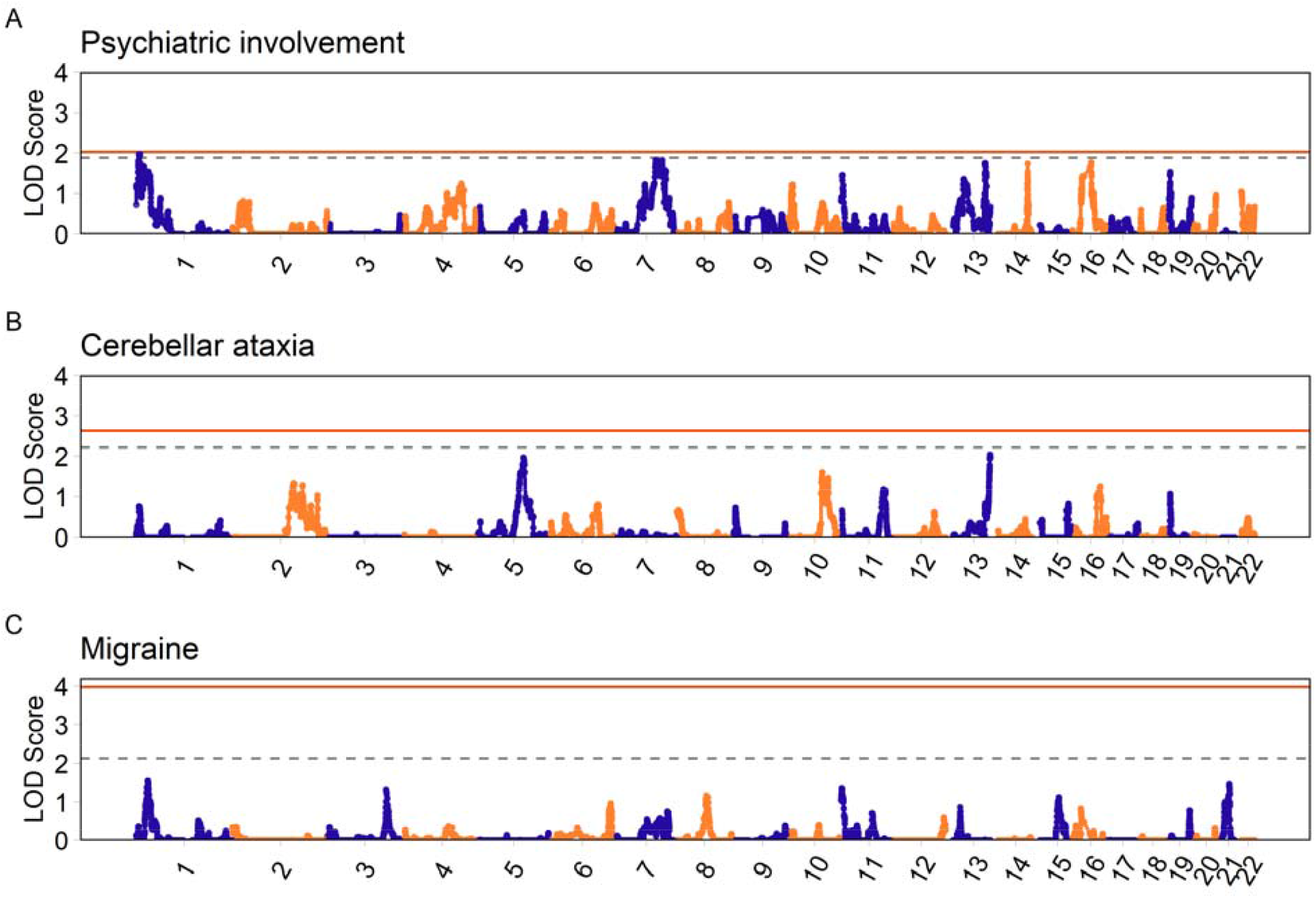
Linkage results for psychiatric involvement, cerebellar ataxia, and migraine. (A) Psychiatric involvement; empirical suggestive LOD ≥ 1.89, empirical LOD significance threshold LOD ≥ 2.03. A region suggestive of genetic linkage was identified on chromosome 1 (1p36.1- 36.11 (LOD_max_=1.97)). Analysis of (B) Cerebellar ataxia (empirical suggestive and significant LOD thresholds ≥ 2.22 and ≥ 2.63 respectively) and (C) migraine (empirical suggestive and significant LOD thresholds ≥ 2.13 and ≥ 3.98 respectively) did not identify suggestive or significant regions of linkage. Empirical LOD score thresholds are shown by the grey dashed (suggestive), and red horizontal (significant) lines.

A summary of the suggestive and significant linkage regions for these neurological phenotypes, their corresponding empirical LOD score thresholds is presented in **Table 3**. Genes encoding mitochondrial proteins (as defined by *MitoCarta3*.*0*^14^) are good candidates within these linked regions, and so we have listed these, although we acknowledge that the causative gene(s) may not be mitochondrial.

**Table 3.**
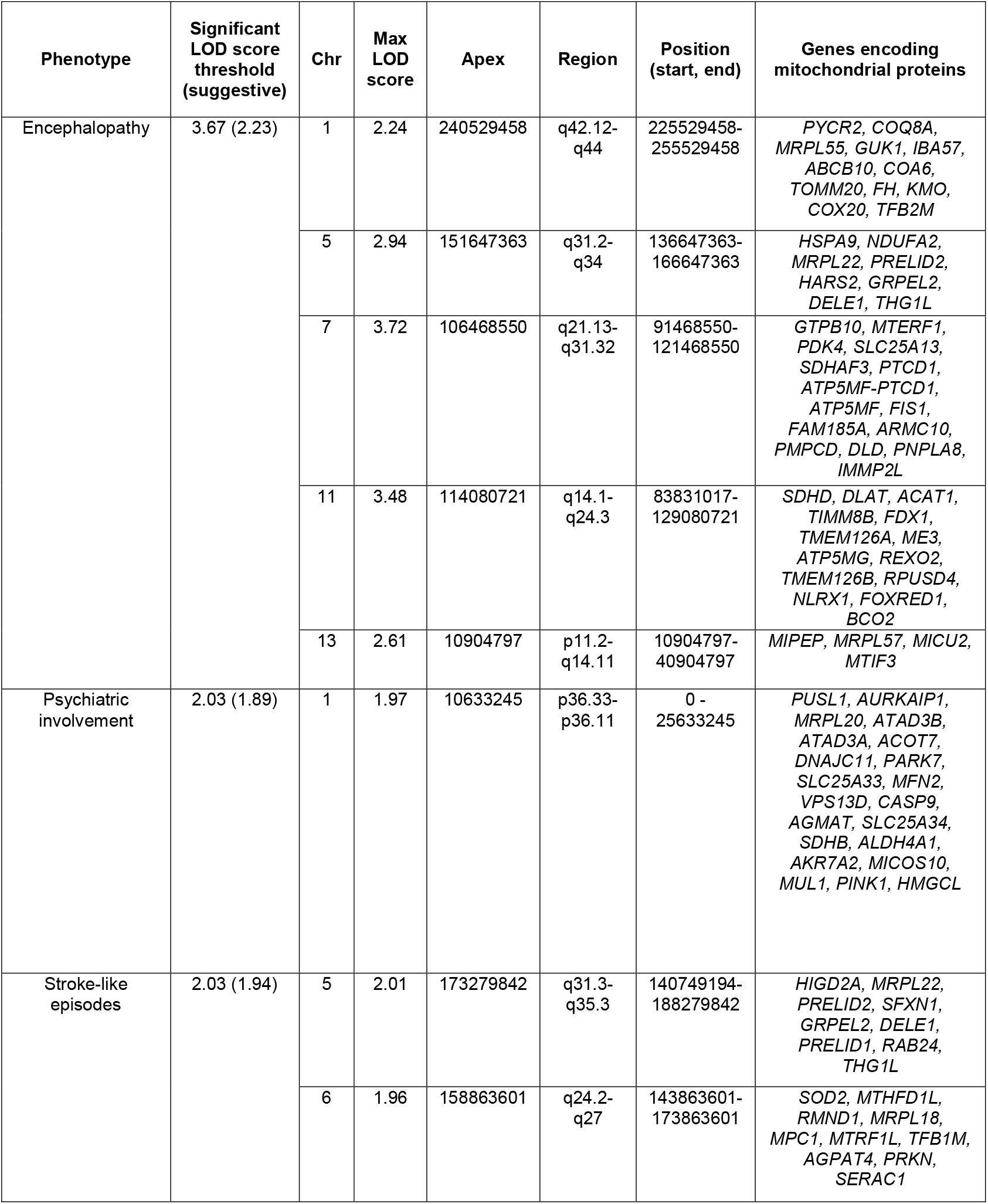
A summary of the suggestive and significant linkage regions identified in the analysis of m.3243A>G-related phenotypes. Genes encoding mitochondrial proteins are defined as those reported in MitoCarta3.0^14^. Positional information relates to GRCh37(hg19) genome build.

### Other neurological phenotypes of m.3243A>G-related disease do not show evidence of genetic linkage

We also considered cerebellar ataxia and migraine, both relatively common neurological phenotypes within our cohort. No regions of suggestive or significant linkage were identified for either phenotype (**Figure 4B and C**). Given the prior evidence of heritability for both phenotypes, the pattern of linkage suggests that the development of these phenotypes may be more polygenic, influenced by a number of additive nuclear genetic factors with a relatively small effect, none of which could be detected through linkage analysis. The null-simulated data for migraine best demonstrates this; the wide range of maximum LOD scores indicates that high LOD scores could be achieved given a large enough genetic effect (**Figure 1**).

### Non-neurological phenotypes of m.3243A>G-related disease

Two of the most common phenotypes of m.3243A>G-related disease are diabetes mellitus and hearing impairment; co-occurrence of these phenotypes is often identified as maternally inherited diabetes and deafness (MIDD), a syndrome with a well-established mitochondrial component^31^. We confirmed the correlation between these phenotypes (r∼0.49, p<0.0001) but no regions of linkage were identified for either phenotype (**Figure 5A and B**). It is possible that the underlying genetic architecture of these phenotypes is more polygenic in structure, which is known to be the case in the wider population in the development of hearing impairment^32,33^ and type II diabetes mellitus^34,35^.

**Figure 5.**
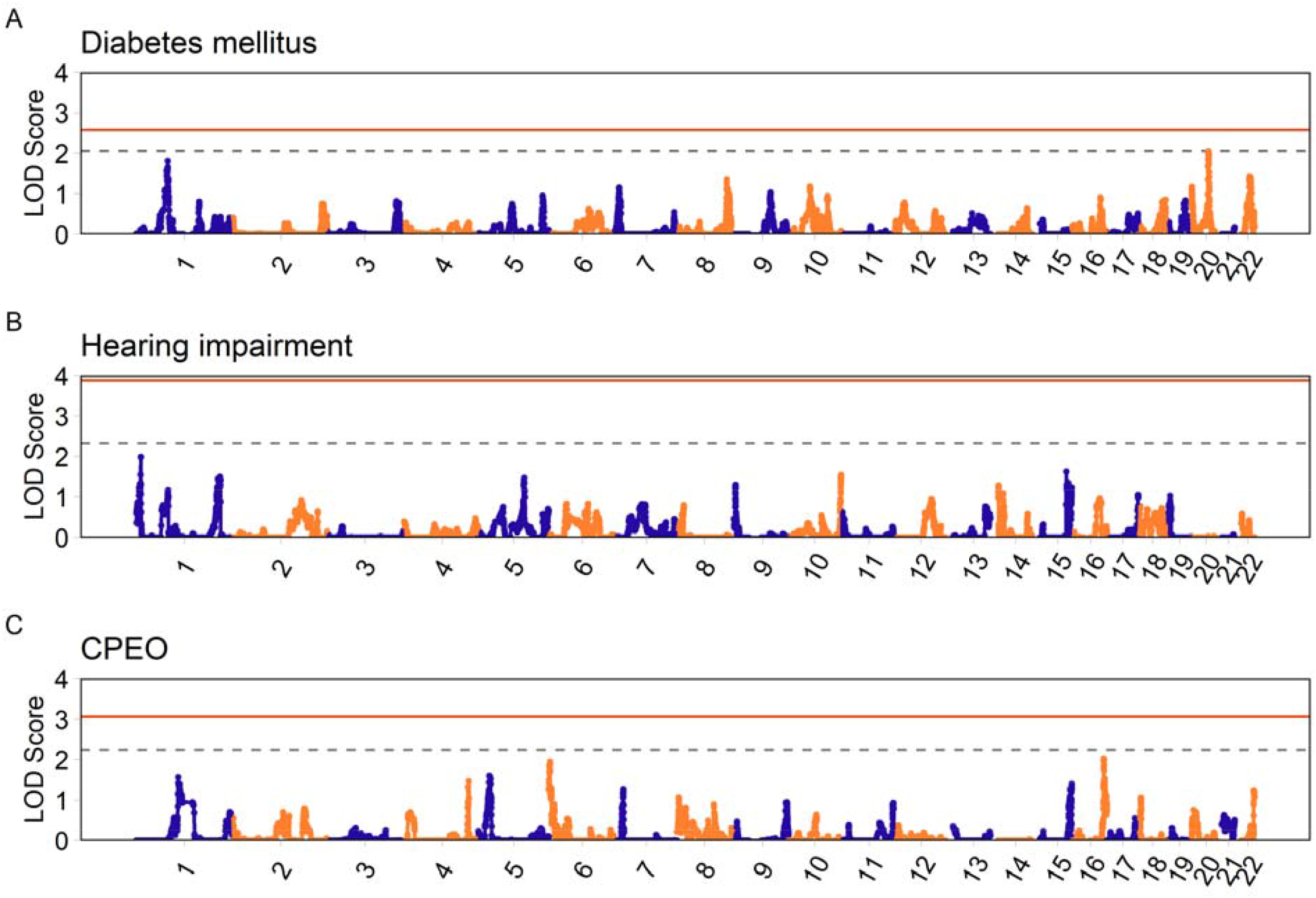
Linkage results for diabetes mellitus, hearing impairment, and CPEO. Using empirical LOD score thresholds there were no regions of suggestive or significant genetic linkage identified in either (A) m.3243A>G-related diabetes (empirical thresholds LOD ≥ 2.06, LOD ≥ 2.53 respectively) (B) hearing impairment (empirical thresholds LOD ≥ 2.32, LOD ≥ 3.88) or (C) CPEO (empirical thresholds LOD ≥ 2.25, LOD ≥ 3.07). Empirical suggestive threshold depicted by dashed grey line; empirical significant threshold depicted by horizontal red line.

We also did not detect any regions of suggestive or significant linkage in the analysis of CPEO, a myopathic phenotype characterised by extraocular muscle weakness (**Figure 5C**). The broad distribution of LODs in the simulation studies (**Figure 1**) suggests that if this phenotype was affected by a small number of nuclear regions with a relatively large effect size, these would have been identifiable through linkage analysis.

## Discussion

Disease pathology caused by pathogenic variants in the mitochondrial genome is clinically heterogeneous, with a large proportion of this heterogeneity likely to be attributable to nuclear genetic variation. By assembling a unique multi-centre collection of m.3243A>G pedigrees and using genetic linkage analysis, we have characterised the nuclear genetic landscape of eight of the most common m.3243A>G-related disease phenotypes. We demonstrated that encephalopathy and stroke-like episodes, severe neurological and cardinal features of MELAS syndrome, are likely to be influenced by a small number of nuclear factors with a relatively large effect size. This is supported by the identification of a significant genetic linkage to encephalopathy on chromosome 7, and suggestive regions for both encephalopathy and stroke-like episodes. This overlap is not surprising given that patients are frequently encephalopathic during the acute presentation of stroke-like episodes.t^36,37^. We also identified a region of suggestive linkage to psychiatric involvement, a feature frequently seen in combination with encephalopathy^12^ but also a feature of the wider group of primary mitochondrial diseases^38^. In contrast, no linkage regions were identified for cerebellar ataxia, migraine, diabetes mellitus, hearing impairment or CPEO, showing that not every disease phenotype is influenced by the nuclear genome in the same manner; any nuclear factors involved in the development of these phenotypes may be more polygenic in nature.

Encephalopathy within m.3243A>G-related disease is highly heritable (h^2^=0.64, SE=0.36, p=0.0408)^12^; the significant linkage region on chromosome 7 is a prime candidate for harbouring factor(s) that influence the development of this severe neurological phenotype. We should not discount any genes lying under the linkage peak but, as genes encoding mitochondrial proteins are strong candidates, we have identified all MitoCarta3.0 genes within this region. Of particular interest is *MTERF1*, encoding Mitochondrial Transcription Termination Factor 1 (mTERF1), an essential mitochondrial transcription termination factor that binds directly to a sequence within *MT-TL1*, encompassing the 3243 position, and has been shown to have a reduced binding efficiency in the presence of m.3243A>G *in vitro*^39–41^. *PMPCB* is another good candidate, encoding the highly-conserved catalytic subunit of mitochondrial pre-sequence protease (MPP), which removes pre-sequences from imported mitochondrial proteins^42^. Bi-allelic variants in *PMPCB* have been found in individuals with early-childhood neurodegeneration, and in some cases epileptic encephalopathy^34^.

Patients presenting with acute stroke-like episodes are frequently encephalopathic; the presence of a shared region of suggestive linkage on chromosome 5 (encephalopathy- LOD_max_=2.94; stroke-like episodes-LOD_max_=2.01) is evidence that these phenotypes may share common nuclear factor(s) that influence their development. This region on chromosome 5 also appears elevated in the analysis of cerebellar ataxia, although it does not surpass the suggestive or significance thresholds, possibly because of a lack of statistical power. However, the correlation of cerebellar ataxia with stroke-like episodes and encephalopathy (r∼0.28-0.35, p<0.0001) suggests that this region may influence all three phenotypes. There have been several nuclear genetic causes of cerebellar ataxia and encephalopathy that have been recognised in the literature; a heterozygous variant in *ATP1A3* (c.2266C >T) was associated with relapsing encephalopathy with cerebellar ataxia (RECA)^43,44^, and presentation of encephalopathy and cerebellar ataxia in Coenzyme Q10 deficiency has been associated with multiple nuclear variants^45^. Bi-allelic autosomal recessive variants in *ADCK3* (also known as *COQ8A*) have also been reported to cause stroke-like episodes and progressive cerebellar ataxia^46^. Although none of these genes are located on chromosome 5, these established cases of one variant causing the simultaneous presentation of these phenotypes furthers the hypothesis that one biological mechanism is responsible for controlling these phenotypes, and one common nuclear modifier could influence the onset of these neurological symptoms. Refining these identified regions will allow further uncovering of the underlying nuclear genetic architecture present in the development of encephalopathy.

We previously established estimates of the heritability (h^2^) of phenotypes of m.324A>G-related disease^10^ which suggested that a large portion of nuclear influence was present in the development of individual m.3243A>G-related phenotypes such as migraine and hearing impairment. Initially, it may seem surprising that we did not detect regions of genetic linkage in these phenotypes, however, the absence of identified linkage regions does not equate to no nuclear involvement, but rather suggests that the underlying architecture of these phenotypes is polygenic in nature, meaning that the influencing power of the nuclear genome is distributed across a lot of common variants with small effect sizes, resembling the pattern of genetic contribution observed in some complex diseases such as diabetes mellitus^34,35^, hearing loss^32,33^, migraine^33^ and psychiatric disturbance^47,48^. It follows that the development of these phenotypes in relation to the m.3243A>G variant may be more complex in nature, involving a combination of environmental factors, which have previously been implicated in some mitochondrial disorders^49–51^, in addition to a polygenic architecture, possibly overlapping with risk for these phenotypes within the general population. Understanding the potential polygenic portion of m.3243A>G-related disease phenotype development may provide insights into disease mechanisms and allow better genetic counselling for patients.

We acknowledge that there are several limitations to our study. Whilst the NMDAS assessment is available for a large number of patients and is clinically validated and useful for tracking the progressive nature of mitochondrial disease in individual patients, the semi-quantitative, ordinal nature of the scale for individual questions presents challenges when it comes to data analysis^21^. To overcome these, and to allow the incorporation of covariates such as m.3243A>G level and age into the analysis, we converted NMDAS scores into binary assessments, however, this means that we have not been able to account for phenotypic severity and may have mis-classified some individuals who were mildly affected at assessment as unaffected. Moreover, It is conceivable that we may underestimate the evolution of multi-system involvement in patients where some clinical features only emerge following the last assessment with NMDAS. On the other hand, some NMDAS questions such as psychiatric disturbance encompass a broad range of conditions which may be associated with the m.3243A>G-related mitochondrial disease incidentally. Furthermore, the NMDAS is a disease assessment tool that can be used in the clinical evaluation of all mitochondrial diseases, therefore, this broader assessment of disease may not capture other manifestations of m.3243A>G-related disease, such as kidney-related disease^52,53^ and macular dystrophy^54^.

Since the first validation and publication of the NMDAS scale in 2006^21^, the understanding, clinical definitions and the diagnostic criteria of some phenotypes have been refined, meaning that the historical information captured through the NMDAS may not accurately reflect the current diagnosis of an individual. This is particularly the case for stroke-like episodes; our NMDAS-derived definition (NMDAS>=1) includes some patients with self-reported “transient focal sensory symptoms” in the preceding 12-month period who may not have had clinical assessment and relevant investigations such as neuroimaging and EEG study as stipulated by the current consensus statement of the diagnostic criteria of stroke-like episodes^36^. Additionally, for this work we have used measures of m.3243A>G-level from routinely and easily accessible patient tissue (blood and urine) which may not reflect the proportion of m.3243A>G present in affected tissues, such as brain.

Despite assembling a uniquely large cohort of m.3243A>G carriers, our cohort is still relatively small compared to genetic linkage study for common, complex disease^15^. Smaller cohorts continue to be an issue when investigating rare disease, presenting challenges to researchers^55^. However, genetic risk factors with large effect sizes can be detected, as recently demonstrated for reversible infantile respiratory chain deficiency, a rare mitochondrial disease caused by variants in both the mitochondrial and nuclear genomes^39^. Importantly, the inability to detect genetic modifiers does not mean that they are not present, indeed, small cohorts combined with genetic heterogeneity have likely hampered past efforts to identify causative nuclear modifiers of mtDNA disease within linkage regions^56,57^. This emphasises the value of the future the assembly of larger, international cohorts to fully understand mtDNA disease heterogeneity.

Understanding disease heterogeneity is a clear priority for mitochondrial disease patients, which is ultimately where research into nuclear modifiers of mtDNA disease will have the biggest, tangible impact^58^. Our results show that the factors likely to have the biggest impact are those that affect severe neurological phenotypes associated with MELAS syndrome, have prepared the way for their future identification and have demonstrated the importance of assembling large well-phenotyped patient cohorts for mtDNA diseases. A deeper understanding of the genetics and molecular mechanisms underlying m.3243A>G-related phenotypes will enable the implementation of personalised medicine and tailored reproductive advice^59,60^ and may identify novel therapeutic targets not only for m.3243A>G-related disease but also for wider mtDNA-related pathologies^61^.

## Supporting information

All supplemental files

## Data Availability

All data available upon request to the authors

## Acknowledgements

We would like to thank the patients within our cohort for participating in this study and the clinical, laboratory and research administration and support teams within all contributing centres. Ethical approval was granted by the Newcastle and North Tyneside Research Ethics Committee (13/NE/0326 and 16/NE/0267), by the Queen Square Research Ethics Committee, London, UK (09/H0716/76), the Ethics Committee of the LMU Munich (182-09), the Ethics committee of the Technical University Munich (200/15 S-SR), the North Wales ethics committee (17/WA/0327) and Italian local Ethical Committees. This research made use of the Rocket High Performance Computing service at Newcastle University.

## Supplementary material

Provided in a separate document

