## Supplementary material for "Defining the nuclear genetic architecture of a common maternally inherited mitochondrial disorder": All supplemental files

| **Phenotype** | **Empirical suggestive LOD threshold** | **Empirical significant LOD threshold** |
| --- | --- | --- |
| **Psychiatric involvement** | 1.89 | 2.03 |
| **Cerebellar ataxia** | 2.22 | 2.63 |
| **Migraine** | 2.13 | 3.98 |
| **Encephalopathy** | 2.23 | 3.67 |
| **Stroke-like episodes** | 1.94 | 2.03 |
| **CPEO** | 2.25 | 3.07 |
| **Hearing impairment** | 2.32 | 3.88 |
| **Diabetes mellitus** | 2.06 | 2.58 |

**Supplementary Table 1.** The calculated empirical suggestive and significant LOD thresholds.

| **Phenotype** | **Total number of individuals with a phenotype assessment** | **Total number of affected individuals** |
| --- | --- | --- |
| **Psychiatric involvement** | 6 | 3 |
| **Cerebellar ataxia** | 6 | 2 |
| **Migraine** | 6 | 1 |
| **Cognition** | 6 | 1 |
| **Neuropathy** | 6 | 2 |
| **Dysphonia-dysarthria** | 6 | 0 |
| **Seizures** | 8 | 2 |
| **Encephalopathy** | 6 | 1 |
| **Stroke-like episodes** | 6 | 0 |
| **CPEO** | 6 | 1 |
| **Hearing Impairment** | 21 | 20 |
| **Gastrointestinal disturbance** | 6 | 1 |
| **Myopathy** | 6 | 2 |
| **Diabetes mellitus** | 115 | 107 |
| **Cardiovascular involvement** | 9 | 6 |

**Supplementary Table 2**. Phenotypic profile of the Exeter sub-cohort. For six individuals, we were able to obtain NMDAS assessments from the NHS Highly Specialised Services for Rare Mitochondrial Disorders, Newcastle upon Tyne as individuals had attended both clinical services. All other phenotypic assessments were available through the Exeter Genomics Laboratory.

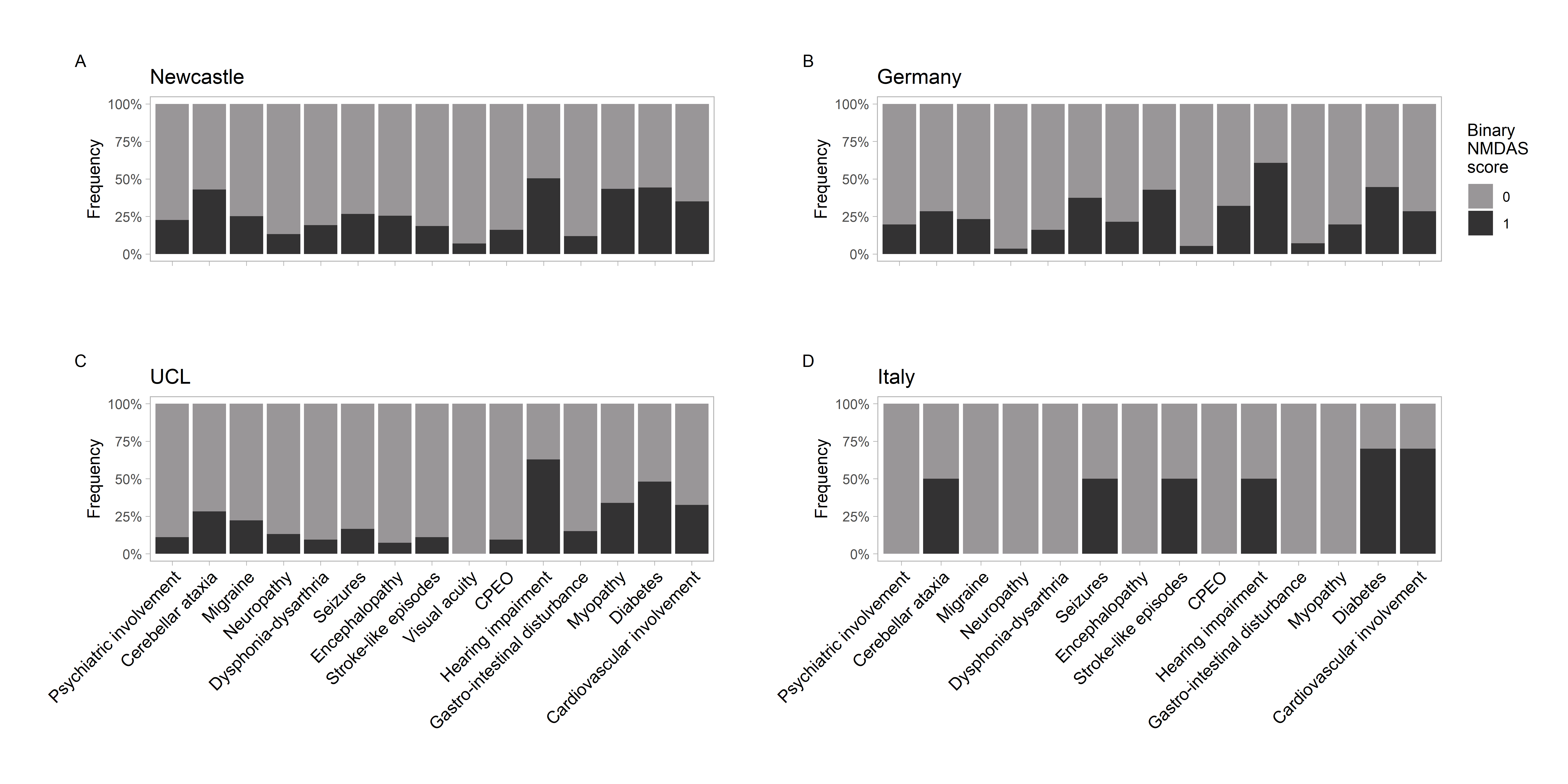

**Supplementary Figure 1.** Binary phenotype distributions within sub-cohorts, as defined in Table 2. (A) Newcastle, UK: n = 231 - 258; (B) Germany: n = 56; (C) UCL, UK: n = 49 – 54, (cognition = 16); (D) Italy n = 10. Cognition was not assessed in either the Germany or Italy sub-cohorts.
